# High Risk Infants who are Low Dose Tolerant after Peanut Oral Food Challenges

**DOI:** 10.1101/2020.01.31.20019570

**Authors:** Adora Lin, Burcin Uygungil, Karen Robbins, Olivia Ackerman, Hemant Sharma

## Abstract

**Background:** Early peanut (PN) introduction may prevent peanut allergy in at-risk children. Little data exists regarding early introduction for infants with large skin prick tests (SPT) or high peanut IgE levels, who are not often offered oral food challenges (OFC).

**Objective:** To retrospectively assess tolerance of a low dose (1 gram) of peanut in infants at risk for peanut allergy, including highest-risk infants (HRI) with SPT wheal >7mm.

**Methods:** We reviewed PN OFCs performed over a two-year period at our pediatric allergy center. Low-dose PN OFC was offered to all infants considered at risk for PN allergy, regardless of peanut SPT or IgE results. Dosing was escalated after OFC at home.

**Results:** Of infants with SPT wheal <=4 mm (n=30), 29 (97%) were low-dose-tolerant. Of those with SPT >4 mm (n=40), 25 (63%) were low-dose-tolerant, and Ara h2 IgE was significantly lower compared to non-tolerant individuals (median 0.62 versus 6.49 kU/L, p<0.05). Among HRI with SPT >7mm (n=22), 12 (55%) were tolerant, with median SPT 9mm (range 8-11mm), PN-IgE 1.1 kU/L (0.3-10.7 kU/L) and Ara h2 1.6 kU/L (0-9.57 kU/L). Age, sex, race, eczema, and egg sensitization did not affect tolerance regardless of SPT size. After 3-6 months, most infants tolerant at OFC were gradually able to consume larger doses of PN.

**Conclusion:** Many infants with PN-SPT >4mm are tolerant of low-dose peanut, and Ara h 2 IgE may be predictive for clinical tolerance among these infants. Low-dose PN-OFC with gradual updosing may help prevent PN allergy in a greater number of at-risk infants.

## Introduction

The landmark Learning Early About Peanut Allergy (LEAP^1^) trial, follow up study (LEAP-On^2^), and other recent data suggest that early peanut (PN) introduction can help to prevent PN allergy in a significant number of at risk infants. Expert panel guidelines have since been published on PN introduction and oral food challenges (OFCs) that allocate risk level and stratify challenges based on who should be challenged at home, in a supervised visit or with an office OFC.^3,4^ Despite these developments and changes in practice, our understanding of the most effective ways to implement early PN introduction is incomplete.

The LEAP trial identified children at high risk for PN allergy in a narrowly defined population (infants with moderate to severe eczema and/or egg allergy) and of those, excluded infants who had skin prick tests over 4 mm. The NIAID consensus guidelines recognize some uncertainty in management of children with larger skin tests, recommending PN introduction or OFC in those who have skin prick tests ≤ 7mm. But these recommendations do not specifically address what the allergy specialist should do with infants who have larger SPTs (or high PN-IgE levels) ^3,4^. Although these recommendations and previous data suggest that those with SPT ≥8 mm are ‘probably allergic’, effectiveness of early PN introduction in young infants with larger SPTs is not known and was not addressed in the LEAP trial. Conceivably many children in this category would benefit from PN-OFC.

To date, published data on implementation of the new PN guidelines in the clinical setting is limited. In a recent retrospective study, the feasibility of implementing these guidelines was addressed in a large referral population of infants under 12 months who were evaluated for PN allergy following the consensus recommendations. Observed feeding or graded food challenges to PN were only offered if SPT <8 mm, while those with larger test results were advised strict avoidance. Reaction rate was low and most were mild in severity^5^.

Both the NIAID guidelines and LEAP trial utilize a goal consumption dose of 3.9 g PN protein for OFC for those with positive testing (LEAP gave single 2 gram PN protein dose to those with negative testing). This corresponds to approximately 1 tablespoon of smooth PN butter or 40 pieces of Bamba, which in our experience, is not feasible for a young infant to reliably consume in one sitting. There are currently no published data on the use of lower-dose PN protein for infant OFCs to assess for low-dose tolerance, though this could potentially allow more successful PN introduction in young infants who are concurrently developing feeding skills and preferences. Taking advantage of low-dose tolerance has been discussed in a clinical communication with older PN allergic children which helped support our clinical reasoning.^6^ Given the major shift in the paradigm for PN introduction after the LEAP trial but prior to the release of the guidelines, we sought to widely offer infant food challenges in a safe setting using low-dose PN-OFC. The “high-risk” infants as described in the LEAP trial excludes many who we deemed, along with the respective families, to be worth challenging given that SPT and PN-IgE levels are not standardized in this age group. Most reference values are based on infants > 12 months^7,8^ although the cutoff of >4mm is referenced for children < 2 years of age.^9^ Because OFC is the gold standard for diagnosis, we considered offering OFCs to the “high-risk” and highest-risk infants an ethically sound service; the potential benefits of tolerating OFC offered in a hospital-based setting with ample emergency resources are great. Based on clinical observations, our approach evolved quickly to using lower PN protein doses (1 gram) for OFCs since this is more age-appropriate in lieu of the almost 4 grams reported in LEAP and standard OFCs. This dose was then slowly increased at home by a set of recommendations to families thought to be more akin to real-world food introduction and escalation. With extensive counseling of families before and after the OFCs, this was considered to be a reasonable off-label clinical service not requiring a formal research protocol.

## Methods

We defined highest-risk infants (HRIs) as those with PN-SPT >7 mm although we offered the challenges to all infants coming to the clinic if home introduction was not desired. Three families at the beginning were offered 2.5 or 5 gram challenges, but most of the HRIs were offered 1 gram as we tailored our approach over time. We have standing IRB approval for OFCs and maintaining a database of these outcomes.

The goal PN protein dose was 1 gram in six gradually escalating doses. Families were referred often for positive IgE testing performed by an outside provider, moderate to severe eczema, egg allergy, possible reaction at first PN introduction, or a sibling with food allergy (FA). Having a food allergic sibling has in some studies been shown to increase risk of FA;^10-12^ thus, we included this in our highest-risk criteria as many of the families were apprehensive about home introduction and delayed introduction was likely to occur.

Informed consent was obtained prior to OFC, in keeping with our standard practices considering OFCs as procedures. The main OFC outcome was the assessment of tolerance to 1 gram of PN protein. PRACTALL guidelines were used to determine if patients did or did not tolerate OFC^13^.

A retrospective chart review was conducted of PN-OFCs performed on 70 infants ages 6-13 months between December 2015 to December 2017 at our pediatric allergy referral center. We compared PN-SPT size, PN-IgE and Ara h 2 IgE between those who were low-dose tolerant and those who had an IgE-mediated reaction during the challenge. There were 3 infants who had delayed reactions including persistent emesis 1.5-3 hours after exposure. These infants were not included in this analysis but represent a previously rare population with PN-FPIES who we have described elsewhere^14^.

If the infant was low-dose PN tolerant, we advised that families introduce 1 gram PN protein (1 teaspoon PN butter, PN powder/flour or 1/4 bag of Bamba) at home 3 times per week and increase by 25% per serving (about 1/4 teaspoon) every 2-4 weeks as tolerated up to a goal of 2 grams of PN 3 times per week up to age 5 years. We discussed with families of children who had the highest testing (SPT > 7mm) that their child is not considered to have tolerance to PN and that they must continue to carry an epinephrine autoinjector at all times until further follow up. Epinephrine autoinjector prescriptions were kept up to date for these families. We also advised that infants be observed closely after home doses for 1-2 hours and that families consider skipping PN on days with fever or gastrointestinal symptoms. We did not consider this to be oral immunotherapy per se since starting doses were much higher and home dosing was not highly regimented.

Statistical analysis was performed in Stata/SE 13.1 using Student’s t test, rank sum, Fisher’s exact analyses and receiver operating characteristic (ROC) area under the curve.

## Results

Over a two-year period, most infants ages 6-13 months at our referral center who were considered high- or highest-risk were low-dose tolerant of a 1 gram PN-OFC (54/70, 77%). 11 patients had successful home introduction and were not challenged (SPT 0-2mm or IgE 0-0.39 kU/L). 6 infants were tested and parents preferred to avoid PN (SPT median 9.5 mm (range 0-12), and median PN-IgE 24.2 kU/L (range 13.8 to >100 kU/L). There were about 8 infants deemed tolerant who had mild, transient, self-resolving hives during the food challenges.

There was no statistical difference in age, gender or race between the groups regardless of SPT size (see Table 1). We had a relatively large percentage of African-Americans compared to LEAP and showed that these infants tolerated PN as often if not more than Caucasians (not significant). This population had predominantly mild atopic dermatitis (AD; 74%) with only about 26% of patients having moderate-to-severe AD determined by clinical judgment by various providers in our group (N.B. we did not have baseline eczema as a reference prior to referral, so eczema might have been more severe). There was no difference in AD, moderate-to-severe AD, or egg sensitization between the groups.

**Table 1.**
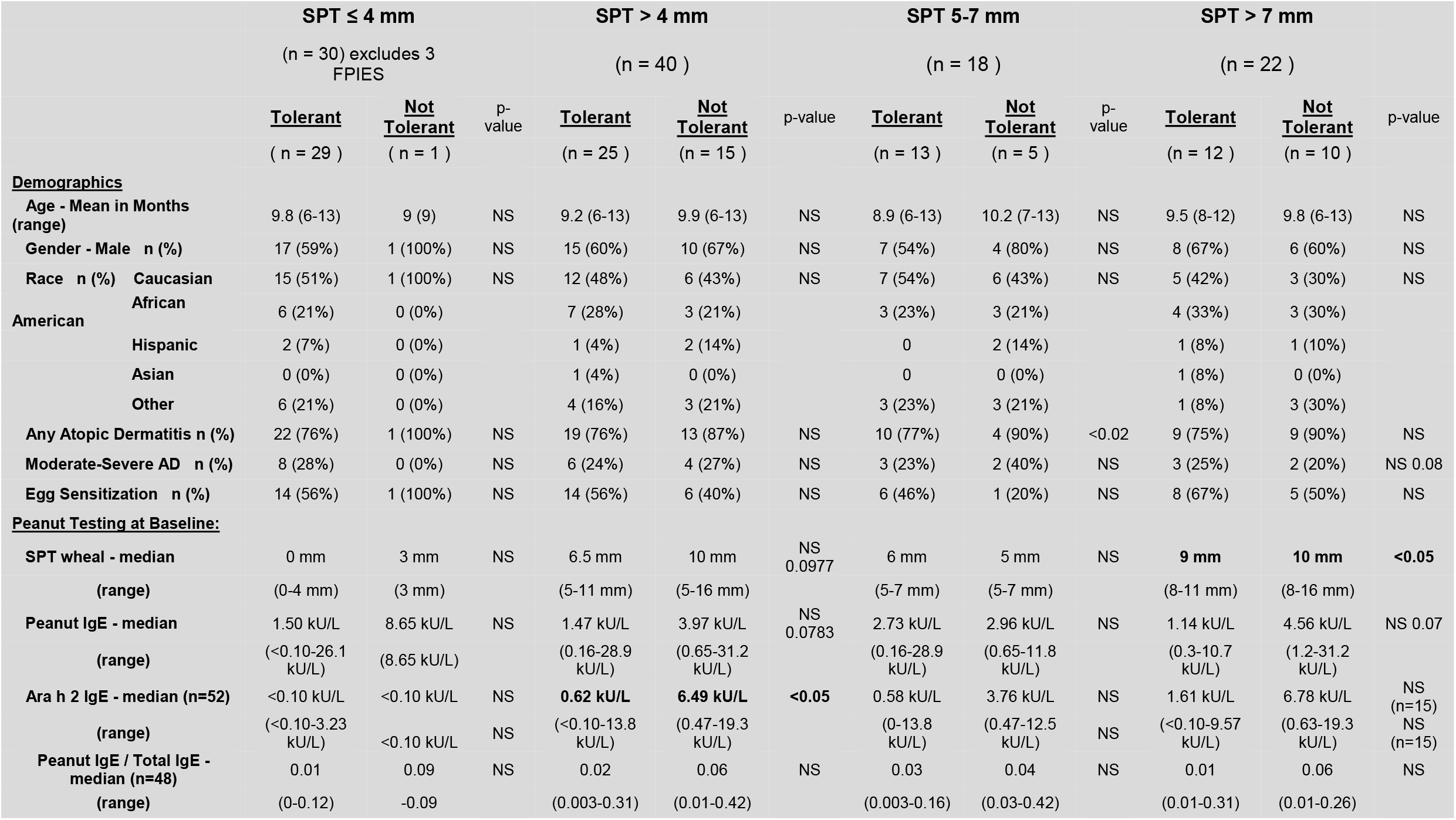
Demographic and biomarker results for PN-OFC by tolerance and SPT size. Statistical analyses were performed in Stata/SE 13.1.

### Biomarkers

We examined SPT size, total PN-IgE, and Ara h 2 IgE because these are laboratory values widely used in clinical practice, although Ara h 2 IgE is missing for some patients due to problems with availability of this test at the beginning of our study. The median SPT size for those who were low-dose PN tolerant was 4 mm compared to 9.5 mm for those who were non-tolerant (Table 1). Of the infants with PN-SPT ≤4 mm nearly all (29/30, 97%) tolerated PN, indicating these infants are likely safe to try PN at home or in an office setting, which falls in line with current guidelines. Those infants with PN-SPT >4 mm mostly tolerated PN as well (25/40, 63%). With a worst-case assumption that those who refused challenges were intolerant then the tolerance rate would be 54% (25/46).

There was considerable overlap in the PN-SPT size and PN-IgE levels between those who were tolerant and not tolerant, however Ara h 2 IgE appeared to help distinguish if an infant would react, especially in those who already had a large SPT >4 mm (median Ara h 2 IgE was 6.49 kU/L in those who reacted vs. 0.62 kU/L in those who were tolerant). Of those infants with SPT >7mm (n=21), tolerant infants (11/21, 52%) had a median SPT 9mm (8-11mm), PN-IgE 1.1 kU/L (range 0.3-10.7 kU/L) and Ara h 2 IgE 1.6 kU/L (range 0-9.57 kU/L). Those who reacted (10/21) had median SPT 10 mm (8-16mm), PN-IgE 4.6 kU/L (1.18-31.2 kU/L), Ara h 2 IgE 6.8 kU/L (0.63-19.3 kU/L). Ara h 2 IgE was the only marker that was significantly different between the two groups (p<0.03). See Figure 1 for a comparison of Ara h 2 IgE by SPT size. ROC curves using data from all patients showed that Ara h2 of 1.05 kU/L or above was associated with a 75% sensitivity and 72% specificity for PN reaction and a level of 6.05 kU/L or greater was associated with 58% sensitivity and 92% specificity (Area under the curve 0.8). If we assume that those who refused OFC were intolerant, the tolerance rate would be 37% (10/27). Age, sex, race, atopic dermatitis and egg sensitization did not affect tolerance regardless of SPT size.

**Figure 1.**
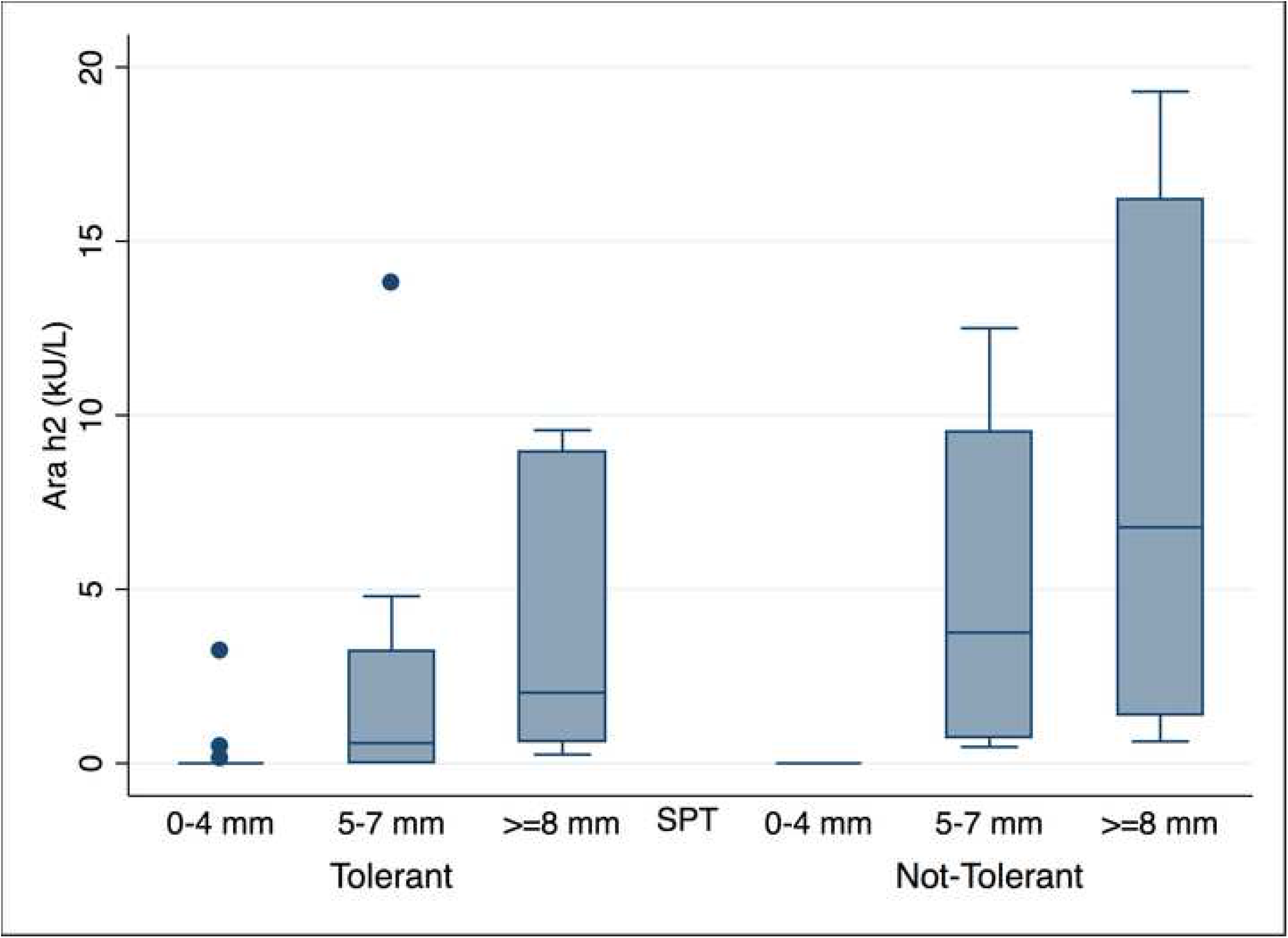
Ara h 2 IgE by SPT size and OFC outcome. Overall, Ara h 2 IgE 1.05 kU/L or above was associated with a 75% sensitivity and 72% specificity for PN reaction and a level of 6.05 kU/L or greater was associated with 58% sensitivity and 92% specificity (Area under the curve 0.8).

There were no biomarkers that could help predict an FPIES reaction in 3 patients. Two of these three patients had moderate to severe eczema, which is higher than the overall population of infants studied here, although clinical and statistical significance was not determined due to the small numbers.

### Safety

Our general practice is to give epinephrine early for reactions during OFCs. Overall, we gave epinephrine to 10 patients who were not PN tolerant with 4 infants receiving 2 doses of epinephrine. All anaphylactic reactions were deemed to be mild based on PRACTALL guidelines. Epinephrine was also given to 2/3 of the FPIES patients prior to determining it was an FPIES reaction, because these were unexpected at the time.

### Follow up

Infants were followed up at a regular clinic visit or by phone if lost to follow up at 3 months, 6 months, and 1 year if possible. Most patients continued with 1 gram of peanut protein 1-3 times per week at minimum and were able to keep it in the diet at home with gradual dose increases as tolerated based on developmental ability. Many of the patients surpassed the goal and are tolerating 4-5 grams of peanut protein 2-3 times per week. One of the infants who initially was not tolerant tried smaller doses of Bamba at home by parental choice and is currently slowly escalating PN at home as desensitization per parental preference. One patient had mild transient hives at home after not eating PN for 2 weeks, but was able to reintroduce PN within 1 week with no further reactions. A few children initially had occasional contact hives with PN ingestion, but no other reactions; parents were comfortable with keeping PN in the diet. There are patients who were deemed to be low-dose tolerant but are no longer eating PN for various reasons including general picky eating and resistance/taste aversion (n=2) or parental misunderstanding of instructions to keep it in the diet (n=1) but these were not associated with higher biomarkers to start.

## Discussion

This study demonstrates successful low-dose PN tolerance in infants who are at high risk for PN allergy including those with the largest PN-SPT (>7 mm). Those who are ≤4 mm might be able to try PN at home or in a more general office setting, given that 97% were tolerant. The highest risk infants in our study would normally be advised to avoid PN based on current guidelines.^3,15-17^ Our PN-OFC results indicate that this would omit many infants who could introduce low-dose PN and possibly avoid a lifelong PN allergy by taking advantage of the theoretical window of opportunity for development of tolerance.^1,2,12,17,18,19^

Potential prevention of PN allergy is arguably the leading advance in food allergy in the last few decades given that the natural history of PN allergy is to persist, especially for children with PN-SPT>13mm or PN-IgE >5 kU/L^20^. Early consumption appears to be critical in many smaller studies^19,21,22^ and was confirmed with a moderately high level of certainty in a meta-analysis based on the LEAP, LEAP-On, and EAT studies^23^. We also know that PN-OIT likely has better outcomes for sustained unresponsiveness in children less than 5 years who already have a known PN allergy^19^. Together, these findings compelled us to try to prevent PN allergy in as many children in our own population as possible.

Given the high impact of prevention and variability of cutoff values based on study site, testing materials, and study population, we offered OFC to our patients as the best way to determine true tolerance. Given the high level of parental anxiety about feeding PN in those who have siblings with FA (n=4) we thought that it was important to include these infants to ensure early introduction even if home introduction was feasible without testing^10-12^. These infants were often referred anyway due to elevated PN-IgE previously obtained by the pediatrician.

We chose to introduce a low dose of 1 gram of PN protein during the PN-OFC instead of 3.9 grams as in LEAP and 5 grams as in standard OFCs. This dose was chosen based on our experience with infants and with PN-OFCs; we have seen that many infants are not easily able to consume more than 1 gram, especially when they are around 6 months of age and have only recently started solid foods. Furthermore the EAT trial demonstrated that it was difficult for families to be adherent given the variation in developmental readiness to eat solid food^18^. For this reason, we chose to challenge infants to a developmentally appropriate dose and increase more gradually at home as a more real world food introduction approach. We strongly recommended that parents give the 1 gram of PN two to three times per week without a long break in consumption based on the LEAP methodology. We also encouraged parents of the highest-risk infants to maintain access to epinephrine autoinjectors until further follow-up.

Based on our data, Ara h 2 IgE seems to be the most valuable test to lab predict reactions during PN-OFCs especially in those who already have a large SPT. PN-SPT and PN-IgE do not appear to be sufficient to exclude an infant from a low-dose OFC. Further studies are required to determine if PN-SPT and Ara h2 in tandem can provide enough assurance that an OFC is not indicated. However, given our current clinical setting within an academic institution with access to emergency and critical care, we feel comfortable offering OFC and managing reactions if the final outcome is to prevent a substantial number of children from becoming PN allergic.

### Study limitations

We consistently evaluate our safety protocols to ensure that all children are being challenged with appropriate resources available. However, we do not expect that the general allergist would be able to offer many of these challenges; thus, access to care will be an issue if this practice becomes more standardized in the future. This was a retrospective study and the standard dose for the OFC changed from the full 3.9 grams as in LEAP to 1 gram, so there were 3 patients initially challenged to higher doses; these patients were deemed tolerant of the low dose. We also did not collect Ara h 2 IgE on all patients as this was a test that became more clinically available and recommended over the course of the study. There was a higher level of mild atopic dermatitis patients in this population as compared to other reports.^1,5^ Atopic dermatitis was based on provider clinical assessment and not a standard score, so it is possible that clinical assessment under-diagnosed moderate to severe eczema, and/or that we did not have access to pre-referral clinical assessments.

### Study strengths

Despite falling outside of guidelines, a number of children demonstrated tolerance to a significant amount of PN protein, allowing PN to be safely incorporated into the diet. To date, little data exists on predictive values of PN-SPT and PN-IgE in infants, and this has been largely due to the fact that these children have not routinely been offered OFCs. While risks exist, this population also stands to benefit significantly from early dietary PN. No severe reactions were noted, no patients required transport to the ED, and none were admitted. In addition to presenting data on a population of children who have not been well studied (large PN SPTs), we also involved an ethnically diverse population, with significant involvement of African American patients (who have been notably underrepresented in previous food allergy research).

### Future goals

We will continue to follow these infants over the next few years and consider enrolling them in studies to examine biomarkers including PN-IgE, Ara h 2 IgE, IgG4/IGE and other immunomodulators. We would like to gather more data on our large African American population (20% of our clinic patients).

## Data Availability

NA

## Acknowledgements

The authors would like to thank Dr. Linda Herbert and Dr. Ashley Ramos for their invaluable assistance with psychological services during the OFCs.

## Abbreviations/acronyms

FA: Food allergy
FPIES: Food protein induced enterocolitis syndrome
HRI: Highest risk infants
LEAP: Learning Early About Peanut Allergy
OFC: Oral food challenge
PN: Peanut
SPT: Skin prick test
AD: atopic dermatitis

## References

1. Du Toit G, Roberts G, Sayre PH, et al. Randomized trial of peanut consumption in infants at risk for peanut allergy. The New England journal of medicine 2015;372:803–13.

2. Du Toit G, Sayre PH, Roberts G, et al. Effect of Avoidance on Peanut Allergy after Early Peanut Consumption. The New England journal of medicine 2016;374:1435–43.

3. Togias A, Cooper SF, Acebal ML, et al. Addendum guidelines for the prevention of peanut allergy in the United States: Report of the National Institute of Allergy and Infectious Diseases-sponsored expert panel. The Journal of allergy and clinical immunology 2017;139:29–44.

4. Bird JA, Groetch M, Allen KJ, et al. Conducting an Oral Food Challenge to Peanut in an Infant. The journal of allergy and clinical immunology In practice 2017;5:301–11 e1.

5. Stukus DR, Prince BT, Mikhail I. Implementation of guidelines for early peanut introduction at a pediatric academic center. The journal of allergy and clinical immunology In practice 2018.

6. Garvey AA, O’Sullivan D, Hourihane JO. Home-based induction of sustained unresponsiveness in children with mild reactions to high doses of peanut. The journal of allergy and clinical immunology In practice 2017;5:1757–9.

7. Dang TD, Tang M, Choo S, et al. Increasing the accuracy of peanut allergy diagnosis by using Ara h 2. The Journal of allergy and clinical immunology 2012;129:1056–63.

8. Roberts G, Lack G. Diagnosing peanut allergy with skin prick and specific IgE testing. The Journal of allergy and clinical immunology 2005;115:1291–6.

9. Hill DJ, Heine RG, Hosking CS. The diagnostic value of skin prick testing in children with food allergy. Pediatric allergy and immunology : official publication of the European Society of Pediatric Allergy and Immunology 2004;15:435–41.

10. Gupta RS, Walkner MM, Greenhawt M, et al. Food Allergy Sensitization and Presentation in Siblings of Food Allergic Children. The journal of allergy and clinical immunology In practice 2016;4:956–62.

11. Begin P, Graham F, Killer K, Paradis J, Paradis L, Des Roches A. Introduction of peanuts in younger siblings of children with peanut allergy: a prospective, double-blinded assessment of risk, of diagnostic tests, and an analysis of patient preferences. Allergy 2016;71:1762–71.

12. Abrams EM, Chan ES, Sicherer SH. Should Younger Siblings of Peanut Allergic Children Be Screened for Peanut Allergy? The journal of allergy and clinical immunology In practice 2018.

13. Sampson HA, Gerth van Wijk R, Bindslev-Jensen C, et al. Standardizing double-blind, placebo-controlled oral food challenges: American Academy of Allergy, Asthma & Immunology-European Academy of Allergy and Clinical Immunology PRACTALL consensus report. The Journal of allergy and clinical immunology 2012;130:1260–74.

14. Robbins KA, Ackerman OR, Carter CA, Uygungil B, Sprunger A, Sharma HP. Food protein-induced enterocolitis syndrome to peanut with early introduction: a clinical dilemma. The journal of allergy and clinical immunology In practice 2017.

15. Sicherer SH, Sampson HA. Food allergy: A review and update on epidemiology, pathogenesis, diagnosis, prevention, and management. The Journal of allergy and clinical immunology 2018;141:41–58.

16. Sampson HA, Aceves S, Bock SA, et al. Food allergy: a practice parameter update-2014. The Journal of allergy and clinical immunology 2014;134:1016–25 e43.

17. Du Toit G, Sampson HA, Plaut M, Burks AW, Akdis CA, Lack G. Food allergy: Update on prevention and tolerance. The Journal of allergy and clinical immunology 2018;141:30–40.

18. Perkin MR, Logan K, Marrs T, et al. Enquiring About Tolerance (EAT) study: Feasibility of an early allergenic food introduction regimen. The Journal of allergy and clinical immunology 2016;137:1477–86 e8.

19. Vickery BP, Berglund JP, Burk CM, et al. Early oral immunotherapy in peanut-allergic preschool children is safe and highly effective. The Journal of allergy and clinical immunology 2017;139:173–81 e8.

20. Begin P, Paradis L, Paradis J, Picard M, Des Roches A. Natural resolution of peanut allergy: a 12-year longitudinal follow-up study. The journal of allergy and clinical immunology In practice 2013;1:528–30 e1-4.

21. Du Toit G, Katz Y, Sasieni P, et al. Early consumption of peanuts in infancy is associated with a low prevalence of peanut allergy. The Journal of allergy and clinical immunology 2008;122:984–91.

22. Fox AT, Sasieni P, du Toit G, Syed H, Lack G. Household peanut consumption as a risk factor for the development of peanut allergy. The Journal of allergy and clinical immunology 2009;123:417–23.

23. Ierodiakonou D, Garcia-Larsen V, Logan A, et al. Timing of Allergenic Food Introduction to the Infant Diet and Risk of Allergic or Autoimmune Disease: A Systematic Review and Meta-analysis. Jama 2016;316:1181–92.

